# A Novel Approach and Apparatus for the Measurement and Evaluation of the Quality of Cardiopulmonary Resuscitation

**DOI:** 10.1101/2023.05.29.23290692

**Authors:** Ishnoor Kaur Bakshi

**Affiliations:** Sidwell Friends School

**Keywords:** Cardiac arrest, cardiopulmonary resuscitation

## Abstract

Cardiac arrest can happen unexpectedly in any place and situation. Cardiopulmonary resuscitation (CPR) is the recommended procedure to be performed in order to maintain the proper blood supply to the brain and prevent death. Cardiopulmonary resuscitation is commonly taught during first-aid courses, but most people do not practice it and therefore might not be able to provide proper assistance to someone suffering cardiac arrest in an emergency. People should be able to practice the procedure on their own in order to learn it properly and be prepared. This means having a feedback system to assess the quality of their training. This research experiment developed and tested a system, capable of assessing the quality of the cardiopulmonary resuscitation procedure and tested it with volunteers with different training and experience and showed that constant practice and evaluation of the procedure significantly improve the ability of the subject to administer it properly.

**Significance Statement:** This research developed and tested a new system to monitor the effectiveness of cardiopulmonary resuscitation during training and practicing for first responders in the case of cardiac arrest.

## I. Introduction

CPR (Cardiopulmonary resuscitation) is intended to restart a heart that has stopped beating, known as cardiac arrest, which is typically caused by an electrical disturbance in the heart muscle. Whatever the cause of cardiac arrest, restarting the heart as quickly as possible to get blood flowing to the brain is essential to preventing permanent brain damage[1]. CPR uses chest compressions to mimic how the heart pumps. These compressions help keep blood flowing throughout the body. Cardiac arrest usually occurs suddenly and often without warning. It is triggered by an electrical malfunction in the heart that causes an irregular heartbeat (arrhythmia). With its pumping action disrupted, the heart cannot pump blood to the brain, lungs and other organs. Seconds later, a person loses consciousness and has no pulse. Death occurs within minutes if the victim does not receive treatment[1]. About 350,000 cardiac arrests happen outside of hospitals each year—and about 7 in 10 of those happen at home[2]. Currently, about 9 in 10 people who have cardiac arrest outside the hospital die[3]. But CPR can help improve those odds. If it is performed in the first few minutes of cardiac arrest, CPR can double or triple a person’s chance of survival[3].

### A. Background research

Provision of CPR has long been the hallmark of cardiac arrest management. Updated evidence from an analysis of over 12500 patients reaffirms the importance of chest compression quality as well as the following: (1) during manual CPR, rescuers should perform chest compressions to a depth of at least 2 inches, or 5 cm, for an average adult while avoiding excessive chest compression depths (greater than 2.4 inches, or 6 cm); (2) It is reasonable for rescuers to perform chest compressions at a rate of 100 to 120 per minute[4]. While CPR performed by professional personnel in hospitals and other healthcare institutions can be considered safe and properly performed, the majority of cardiac arrests occur suddenly in public places or at home. Therefore, the ability of bystanders to perform proper CPR is essential to increase the probability of the person(s) with cardiac arrest to survive and especially to survive without any permanent brain damage. Various studies show that in out-of-home cardiac arrest (OHCA), bystanders in the US attempt CPR in between 14%[5] and 45%[6] of the time, with a median of 32%[7]. CPR is likely to be effective only if commenced within 6 minutes after the blood flow stops[8] because permanent brain cell damage occurs when fresh blood infuses the cells after that time, since the cells of the brain become dormant in as little as 4–6 minutes in an oxygen deprived environment and, therefore, cannot survive the reintroduction of oxygen in a traditional resuscitation. A CARES 2019 study showed that laypeople in the United States initiated CPR in 41.6% of OHCAs[9]. A prospective data collection concerning 10682 OHCA cases from 27 European countries in October 2014 found an incidence of 84 per 100000 people, with CPR attempted in 19 to 104 cases per 100000 people. Return of pulse occurred in 28.6% (range for countries, 9%–50%), with 10.3% (range, 1.1%–30.8%) of people on whom CPR was attempted surviving to hospital discharge or 30 days[9].

### B. Hypothesis

CPR is taught around the world during standard first aid training courses that usually last a couple of days and where a single instructor, sometime a volunteer with a lot of training but limited field practice, needs to oversee the training of several people at the same time. After this two-days course the attendants receive a certification that needs to be renewed every two years by attending a similar course again. Without any constant and recurrent practice, the quality of such training is therefore not optimal. Effective CPR requires a proper rate of compressions per minute as well as sufficient compression depth and pressure. A combination of self-instruction and instructor-led teaching with hands-on training is recommended as an alternative to only instructor-led courses for lay rescuers. If instructor-led training is not available, self-directed training is recommended for lay rescuers[4]. The primary goal of resuscitation training for lay rescuers (i.e., non–healthcare professionals) is to increase immediate bystander CPR rates, automated external defibrillator (AED) use, and timely emergency response system activation during an OHCA[4]. Studies comparing self-instruction or video-based instruction with instructor-led training demonstrate no significant differences in performance outcomes[4]. A shift to more self-directed training may lead to a higher proportion of trained lay rescuers, thus increasing the chances that a trained lay rescuer will be available during OHCA[4].

In order to improve the chances that bystanders will perform effective CPR in case of sudden cardiac arrest it is therefore necessary to provide proper, consistent training and the possibility of self-practice with quantifiable feedback of the CPR performance. In the work here presented we equipped a CPR manikin with different sensors to measure the quality of CPR administered by people with different levels of training and practice. This experiment measured certain metrics (CPM, Compression depth, CPR Cycles) to determine the quality and effectiveness of CPR.

## II. Materials and Methods

### A. Equipment components

The research experiment required assembly and coding of the following equipment. Namely, Arduino UNO controller, breadboard, connection wires, pressure sensors, push button switch, Arduino sketch for coding, laptop for data collection and CPR manikin. **Figure 1** shows the setup clearly. As seen in **Figure 1**, the system includes two different sensors mounted on the manikin, one to measure the pressure and frequency of chest compressions and another to measure the depth of such compressions. The sensors are connected to an Arduino controller paired to a laptop for programming and data collection and recording. The code is hosted at the following GitHub [repo](https://github.com/ishnoor17/CPR_Quality/blob/69c33f6155730283c0193218f17c53036ee3efa4/CPR_Project%20(1).txt). System assembly

**Figure 1.**
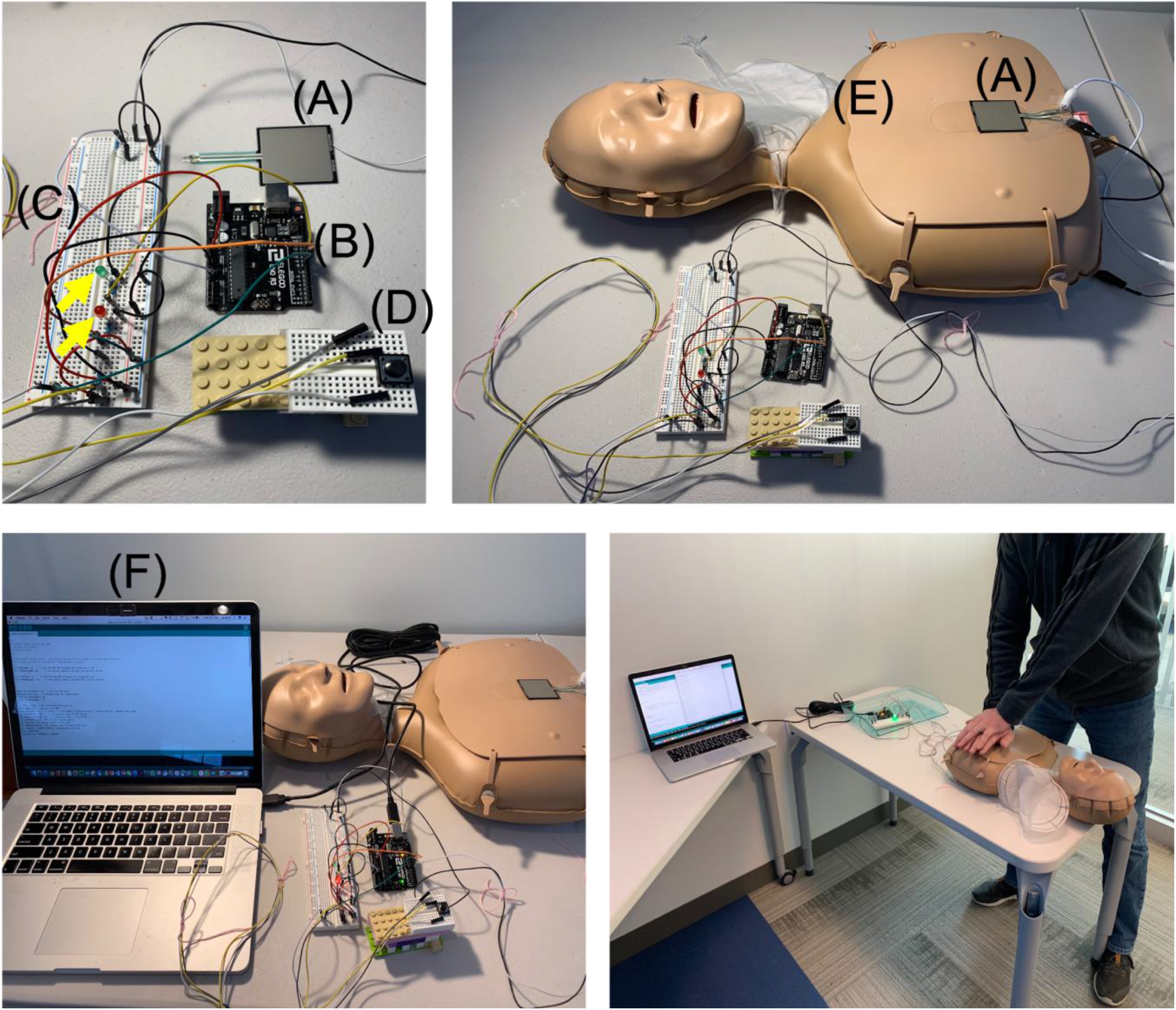
Description of the different components of the system used to study the quality of CPR. Top left photo: (A) pressure sensor, (B) Arduino UNO controller, (C) bread board with indicator LEDs (yello arrows), (D) push button switch. Top right photo: (A) pressure sensor, (E) CPR manikin. Bottom left photo: (F) computer for coding and data collection. Bottom right photo: representative settings of how the data was recorded during a CPR session with one of the study volunteers.

### B. Controller script

The code is hosted at the following GitHub [repo](https://github.com/ishnoor17/CPR_Quality/blob/69c33f6155730283c0193218f17c53036ee3efa4/CPR_Project%20(1).txt).

### C. Procedure

The experiment procedure was as follows: Gather research on CPR procedure and metrics, assemble sensors/switch and Arduino components, develop code, measure data from all sensors, take trials based on location, for each trial collect data, analyze data based on baseline, compare baseline to trials and make conclusions and next steps.

People with different training and experience in providing CPR were recruited on a volunteer basis by the author through personal contacts as well as recommendations. Five different people were recruited for this study representing a continuum from completely unexperienced to professionals administering CPR several times a day in their line of work. All the volunteers agreed to have their data recorded and analysed in the present study and the authors are maintaining their anonymity as concern to their privacy.

There were total of 5×10 trials and 1 baseline trial. Trial 1 was done by a local friend (with little training in CPR), Trial 2 was done by me, (CPR certified), Trial 3 was done by school teacher, (experience and CPR Certified), Trial 4 was done by a Physician, MD (CPR Certified and practitioner), and Trials 5 – Level 2 EMT (Senior CPR Specialist).

### D. Data collection

The instrumentation used in this study was designed and programmed to record data regarding the CPR compressions per minute, a depth of compression of 5 cm (2 inches) and the average compression pressure. The subjects were requested to complete ten consecutive cycles of CPR in a single session.

### E. Data analysis

In order to compare the quality of CPR between the different subjects, the data collected in the present study was analysed to compute the mean values over the ten cycles for each parameter measured as well as the coefficient of variation (defined as the ratio between the standard deviation and the mean value) expressed as percentage.

## III. Results

### A. Approach of experiment

Effective CPR relies on proper technique that needs to be learned and practiced in order to be applied when needed, especially by non-professional bystanders in cases of OHCA. The current methods of teaching CPR include 2-days-long courses taught by volunteers and using passive manikins for practice. Outside of these classes, most people do no practice any further. In order to verify whether trained laypeople can provide effective CPR after a standard CPR course, or even trained but non-practicing people can do so compared to specialists, we set out to outfit a standard CPR manikin with a set of sensors capable to measure the different parameters that define proper CPR administration. As seen in ***Figure 1***, the system includes two different sensors mounted on the manikin, one to measure the pressure and frequency of chest compressions and another to measure the depth of such compressions. The sensors are connected to an Arduino controller paired to a laptop for programming and data collection and recording (see *Materials and Methods* for details).

### B. Trials details and data

We asked five independent subjects, having different levels of training and experience with CPR, to perform CPR using our system. We were able to register the pressure and depth of each compression and the compression frequency. The subjects were asked to complete 10 consecutive cycles of 30 compressions each. The first subject was an adult with no previous training nor experience with CPR; the subject was instructed about the different CPR parameters (compression depth of 5 cm and frequency of 100-120 compressions per minute) just before the test. As reported in ***Figure 2***, the subject was able to maintain a proper compression frequency of about 105 compressions per minute and a proper depth for the first two cycles but then both frequency and depth were not high enough to comply with CPR guidelines. The compression pressure was also highly variable though the test and decreased though the latest cycles. The subject reported that it was physically challenging to maintain proper and constant rate and pressure for the CPR. The second subject had previous training for CPR but did not have any experience outside the training. As reported in ***Figure 2***, the second subject was able to maintain a proper rate of compressions per minute through most of the test even though there was a wide variability between the cycles. However, the subject was not able to administer proper compression depth and the pressure applied decreased toward the ending of the test, mainly due to physical fatigue.

**Figure 2.**
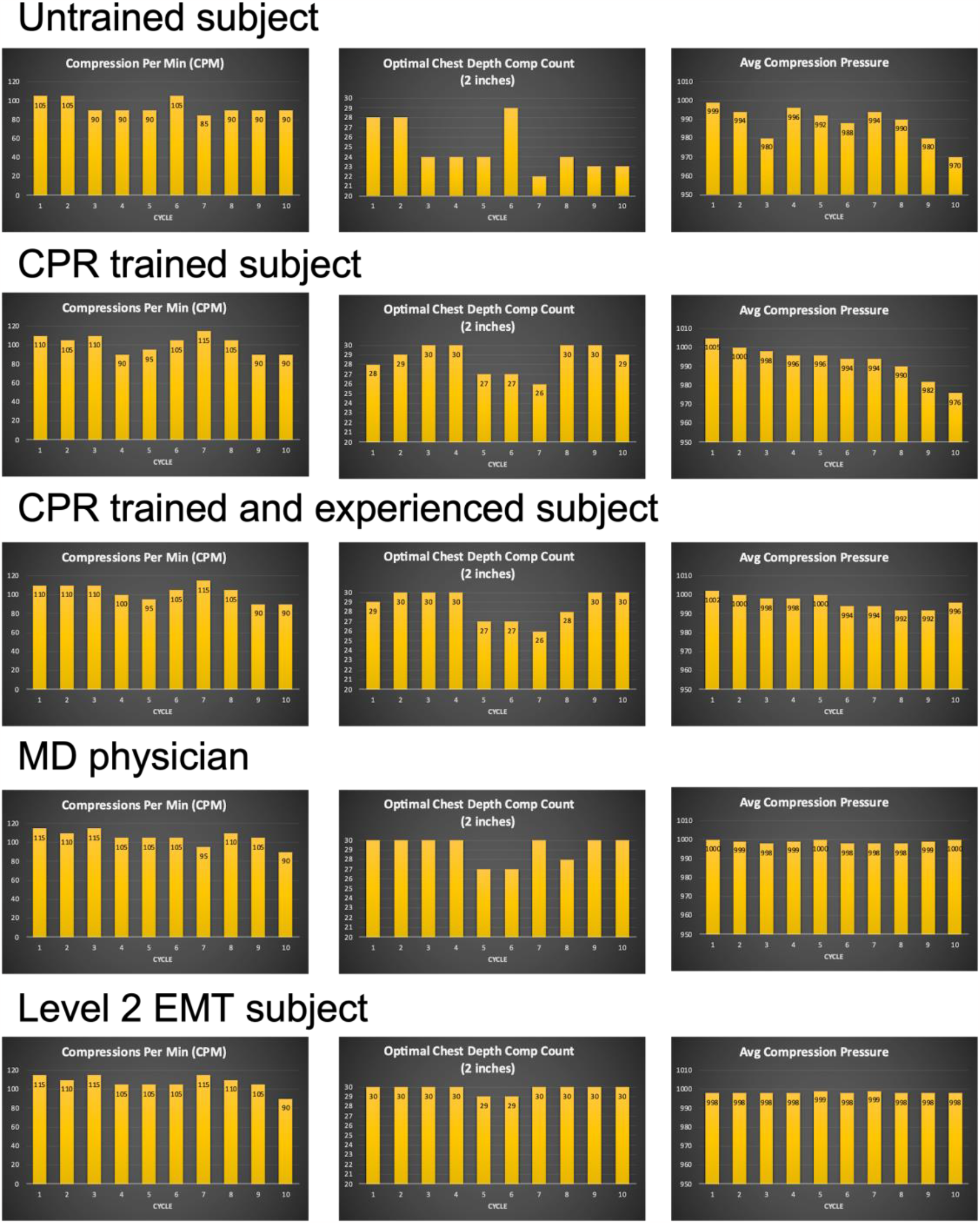
CPR data collected from different operators with different experience in CPR. Different operators used the system described in **Figure 1**. CPR compressions per minute, depth of compression and average compression pressure were recorded for ten consecutive cycles of CPR.

The third subject had previous training in CPR and had some practice in the technique but was not a healthcare worker and did not administer CPR as a specialist. As seen in ***Figure 2***, this subject had similar results as the second subject, even though compression pressure was more constant throughout the test and did not significantly decrease, showing more physical endurance. The thirst subject, a healthcare professional administering CPR in the line of work several times a week, was able to maintain proper compression rate, as well as depth and pressure though most of the cycles of the test (***Figure 2***).

The final subject, a level two emergency medical technician (EMT) administering CPR several times a day, showed the best results recorded in this study. This subject recorded almost perfect results, falling short of compressions per minutes only in the last cycle and providing perfect depth of each compression except in two cycles, which 29 perfect compressions out of 30. Remarkably, the last subject maintained a constant compression pressure throughout the 10 cycles of the test.

We also analysed the recorded data by computing the average compression rate, compression depth and pressure for each subject as well as the coefficient of variation for each parameter, in order to better compare the results between the different subject (***Figure 3***). It is quite clear that the more experienced a subject is, the better he or she can administer effective CPR, defined as CPR complying with international guidelines[4]. Even though some training seemed enough to maintain a proper compression rate though most of the test (***Figure 3***), it is clear that constant practice is required in order to provide proper depth and pressure through the multiples cycles. Considering that, in case of cardiac arrest, bystanders are required to keep administering CPR until professional responders can intervene, people trained in administering CPR should undergo constant practice to be able to perform the procedure properly. This is supported by the results recorded by the fourth and fifth subjects in this study, who administer CPR on a daily or weekly basis and have constant feedback on the effectiveness of their procedure since they are using CPR on patients suffering from cardiac arrest.

**Figure 3.**
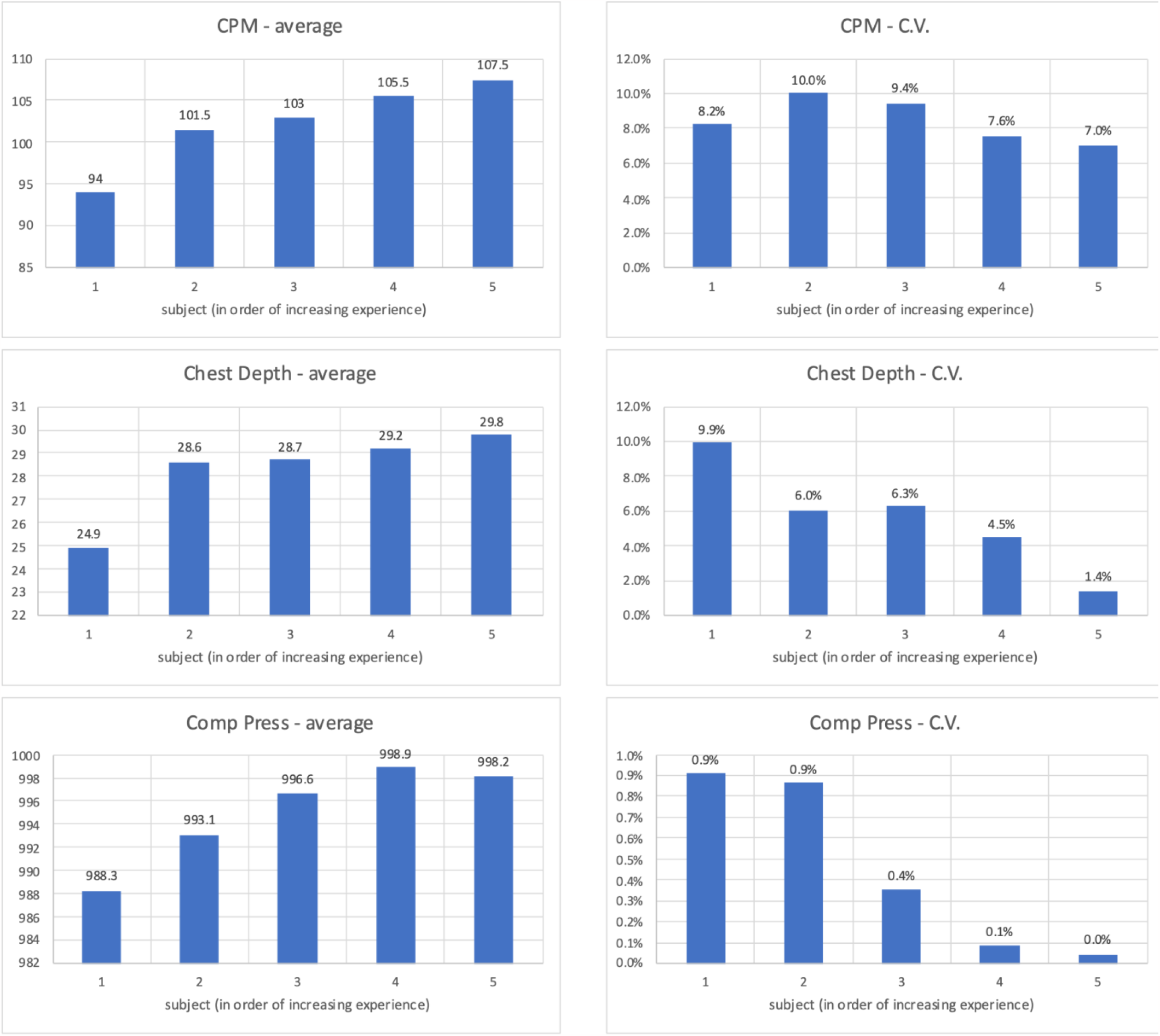
Statistical analysis of the data collected from subject with different experience in performing CPR. The data shown in **Figure 2** was analyzed to evaluate the difference between the different subjects that performed CPR during this study using the system described in Fig. 1. The results from the different subjects were compared using the average and the coefficient of variation (C.V.) of the different parameters recorded over ten consecutive cycles of CPR.

## IV. Discussion

### A. Trial observations

Several decades of field studies have shown that an effective CPR requires a proper rate of compression, between 100 and 120 compressions per minutes, a proper compression depth, at least 5 cm (2 inches) but not more than 6 cm (2.4 inches) performed in cycles of 30 compressions repeated as long as the person with cardiac arrest start breathing again on their own. Emergency support personnel performs CPR procedures several times every day in their line of duty, and therefore this constant forced practice cause their performance to be constantly honed and be as close as the guidelines require. This level of accuracy was recorded in this study using the equipment described by the level 2 EMT subject (***Figure 2***). Medical doctors with years of training, even though with less practice than an EMT specialist, are of course able to perform effective CPR, even though our results show that the less practice generates a bit more of variation from the guidelines (***Figure 3***).

While trained but less experienced subjects seem to be still able to perform CPR of acceptable quality, it starts to become obvious that the lack of practice has a considerable effect on the consistency of the performance. Indeed, trained lay people showed a much bigger variation in the compression rate as well as in the depth of the compressions (***Figure 2*** and ***Figure 3***). The pressure to be provided and the high rate per minute that needs to be maintained in order to provide effective CPR are physically demanding for the performer. This is evident in the results of the recording by subjects one and two in this study (***Figure 2***). Therefore, people with less training and practice are unlikely to be able to perform quality CPR for an appropriate amount of time, enough to allow for professional support to arrive and take over assisting the person with cardiac arrest. The opportunity to self-train with qualitative feedback even outside of first aid curses is therefore of principal importance to make sure that bystanders are able to perform appropriate quality CPR in the case of sudden OHCA.

### B. Conclusions

Effective CPR requires precision and consistency in the compression per minute, compression depth and pressure as showed by the data produced by the EMT professional in this study. It is therefore pivotal that people involved in administering CPR have the means to measure their skills during training. The instrumentation here described has proved to be able to provide these measurements guaranteeing precision and reliability. Such instrumentation could be deployed during in person classes as well as during remote or individual training and practicing. Especially when an experienced instructor cannot be present in person to oversee the trainee, this instrumentation can support as well as validate the correctness of the training.

### C. Other applications in the industry

The prototype presented in this study represents a first approach to the problem of training people of the public to administer effective CPR. The instrumentation described could be easily redesigned to be incorporated in standard manikins that could be used for CPR training and practice by single people without the assistance of experienced instructors. It is possible to envision even a more advanced system that would be able to record and assess the quality of artificial ventilation, that is the second part of the CPR procedure not covered in the study here presented.

## Data Availability

All data produced in the present study are available upon reasonable request to the authors
All data produced in the present work are contained in the manuscript

## Acknowledgements

I want to thank you the participants of this study for volunteering to have their CPR skills recorded and for the time they donated to this study. I want also thank Dr. Singh, MD Emergency Medicine and Dr. Khurana, MD Cardiologist for their support and advice that were used as guidance in the inception of this project.

## Notes

### Competing Interest Statement

The authors have declared no competing interest.

### Funding Statement

This study did not receive any funding

